# Measuring environmental exposures in people’s activity space: The need to account for travel modes and exposure decay

**DOI:** 10.1101/2023.01.06.23284161

**Authors:** Lai Wei, Mei-Po Kwan, Roel Vermeulen, Marco Helbich

## Abstract

**Background:** Accurately quantifying people’s out-of-home environmental exposure is important for identifying disease risk factors. Several activity space-based exposure assessments exist, possibly leading to different exposure estimates, and have neither considered individual travel modes nor exposure-related distance decay effects.

**Objective:** We aimed 1) to develop an activity space-based exposure assessment approach that included travel modes and exposure-related distance decay effects and 2) to compare the size of such spaces and the exposure estimates derived from them across typically used activity space operationalizations.

**Methods:** We used 7-day-long global positioning system (GPS)-enabled smartphone-based tracking data of 269 Dutch adults. People’s GPS trajectory points were classified into passive and active travel modes. Exposure-related distance decay effects were modeled through linear, exponential, and Gaussian decay functions. We performed cross-comparisons on these three functional decay models and an unweighted model in conjunction with four activity space models (i.e., home-based buffers, minimum convex polygons, two standard deviational ellipses, and time-weighted GPS-based buffers). We applied non-parametric Kruskal-Wallis tests, pair-wise Wilcoxon signed-rank tests, and Spearman correlations to assess mean differences in the extent of the activity spaces and correlations across exposures to particulate matter (PM_2.5_), noise, green space, and blue space.

**Results:** Participants spent, on average, 42% of their daily life out-of-home. We observed that including travel modes into activity space delineation resulted in significantly more compact activity spaces. Exposure estimates for PM_2.5_ and blue space were significantly (*p*<0.05) different between exposure estimates that did or did not account for travel modes, unlike noise and green space, for which differences did not reach significance. While the inclusion of distance decay effects significantly affected noise and green space exposure assessments, the decay functions applied appear not to have had any impact on the results. We found that residential exposure estimates appear appropriate for use as proxy values for the overall amount of PM_2.5_ exposure in people’s daily lives, while GPS-based assessments are suitable for noise, green space, and blue space.

**Significance:** For some exposures, the tested activity space definitions, although significantly correlated, exhibited differing exposure estimate results based on inclusion or exclusion of travel modes or distance decay effect. Results only supported using home-based buffer values as proxies for individuals’ daily short-term PM_2.5_ exposure.

**Impact statement:** Accurately quantifying people’s out-of-home environmental exposure is vital for identifying disease risk and protective factors. Although many activity space-based exposure assessments exist, these approaches possibly lead to different exposure estimates. We methodologically and conceptually innovate by developing an activity space-based exposure assessment considering people’s travel modes and exposure-related distance decay effect. Our comparison with other activity spaces provides novel insights into dynamic exposure assessment approaches. Despite most epidemiological studies still considering people’s homes as the sole exposure location, our study is fundamental because people are typically exposed to multiple out-of-home environmental contexts.

## 1 Introduction

There is growing recognition of the importance of environmental exposure to human health ^1^. Many meta-analyses and reviews have reported suggestive associations between people’s physical and mental health and exposure to blue space ^2^, green space ^3^, air pollution ^4^, and noise ^5^. Yet, conflicting results were often reported across studies, possibly due to the inconsistent measurement and operationalization of environmental exposures.

Most previous studies solely assessed exposures based on people’s home locations ^6, 7^. Administrative units and circular or network buffers centered on people’s home addresses have been widely adopted for use in the estimation of people’s exposures to the environment. However, these approaches focus solely on home locations and disregard people’s day-to-day mobility, which frequently extends beyond the residential neighborhood ^8, 9^.

Individuals’ daily lives typically comprise multiple activity places (e.g., work), typically situated beyond the home location ^10^. These places a person routinely visits are conceptualized as ‘activity spaces’ ^11^. As elucidated by the uncertain geographic context problem ^9^, the residential location possibly only accounts for a (small) proportion of human activity spaces ^12^. Consequently, using only individuals’ residential neighborhoods to measure their environmental exposure, as typically done in epidemiological studies ^13–15^, may yield inaccurate measurements of actual exposures, affecting estimates of health-environment associations ^8, 16^.

Activity space-based exposure assessments, grounding on time geography ^17^, have been proposed as a possible solution to the uncertain geographic context problem ^18–20^. This solution has been supported by the development and application of portable devices equipped with global positioning systems (GPS), allowing people’s daily mobility to be captured with finer space-time granularity than traditional data collection approaches based on questionnaires and travel and activity diaries ^21, 22^.

A growing number of studies have assessed exposures along individuals’ mobility paths using GPS-based locational data ^23–28^. Typically, these studies have implemented buffers centered on each GPS point or along people’s moving trajectories as exposure receptor areas ^20, 23^. Due to a lack of any standard widely accepted by scholars for buffer size selection, the selection of buffer sizes and shapes was typically determined ad hoc ^29^. Furthermore, fewer studies have also included exposure time-weightings to capture dwell-time ^26, 30–32^. Despite the progress made in previous exposure assessment studies using locational data, two limitations based on previous approaches remain.

First, neglecting individuals’ unique combinations of activity-travel characteristics could lead to inaccurate exposure assessments ^31^. Individuals’ exposure levels are associated with different modes of transportation and the settings of the traversed environment ^29, 33, 34^. For instance, people are more isolated from their surroundings and immersed in the transport-related micro-environment when traveling in a vehicle rather than walking or cycling ^35, 36^. However, previous studies primarily explored how active travel mode affects exposure assessment ^29, 37^, with passive travel mode receiving less attention.

Second, exposure-related distance decay effects have been neglected in the past. Assessments in current studies assume that environmental exposures within pre-defined buffers (typically intra-buffer mean exposure values) have similar health effects ^24^. However, it is reasonable to assume that health effects may vary with distance; i.e., environmental risks closer to individuals may substantially impact their health more than those more distant ^38–40^. If distance decay effects are omitted, more distant exposures with potentially less influence on health could be overestimated, particularly as buffer sizes increase. Previously, although conceptually sound ^19, 41^, distance decay functions were rarely adopted in environmental exposure studies.

To address the identified knowledge gaps, this study 1) developed an activity space-based exposure assessment approach that includes individuals’ travel modes and exposure-related distance decay effects and 2) compared multiple environmental exposures across different activity space definitions. Our first hypothesis was that including travel modes would reduce the contextual unit’s extent and also increase the level of exposure captured. Furthermore, our second hypothesis expected that incorporating distance decay into the exposure assessments would lead to lower observed exposure levels.

## 2 Materials and methods

### 2.1 Data collection

We gathered cross-sectional tracking data from the Netherlands ^42^. People’s locational data were collected through GPS-enabled smartphones between September and November 2018. Data collection followed a two-step procedure. First, 45,000 people recruited using stratified random sampling from the Dutch population aged 18-65 were invited to conduct an online survey. Second, those respondents who agreed to be re-contacted were asked to consent to be locationally tracked via a smartphone-based application. The application was developed for Android operating system versions ≥4.4. From an initial 8,869 invitations, 820 invitees downloaded the app, and of those, 753 became participants by agreeing to allow the research team to collect smartphone sensor data. The need for user interaction was kept to a minimum to reduce the risk of behavioral changes on the part of participants as a result of being GPS-tracked.

Given the smartphone brands participants possessed and their concomitant battery life, the application was designed to use an adaptive sampling strategy for collecting locational data. The default time interval to record a participant’s location was every 20 seconds. However, after 30 minutes of lack of movement (<20 m), the interval was adjusted to once per minute and after 60 minutes, to once every two minutes. After the app recorded a cumulative total of 7 days of data, it terminated data acquisition. Daily data uploads were stored on a secure server at Utrecht University. The study protocol was approved by the Ethics Review Board of Utrecht University (FETC17-060).

### 2.2 Approaches for context delineation

We cross-compared a static exposure assessment approach (i.e., home-based buffers), three typical activity space-based approaches, and our refined GPS-based approach for contextual unit^1^ delineation.

#### 2.2.1 Home-based buffers

The home-based buffers (HB) were operationalized through circular exposure windows with 1 km radii centered at the home location (see supplementary Fig. S1) ^20^. We inferred people’s home locations by dwell time length, with the locations having the most extended dwell times (across the tracking duration) used as proxies for their home addresses ^20^. Exposures were calculated by averaging raster cell values of the exposure surfaces within the buffer.

#### 2.2.2 Typical activity space-based approaches

As used elsewhere ^20, 28^, we implemented three activity-based approaches (Fig. S1). First, the minimum convex polygon (MCP) refers to the smallest convex polygon, including all mobility paths an individual uses daily. We aggregated these daily MCPs per participant over their 7-day tracking periods. Second, the standard deviational ellipse (SDE) describes the directional distribution and dispersion of a set of GPS locations. We used an SDE based on two standard deviations covering approximately 95% of a participant’s GPS points ^43^. SDEs were generated and merged for each participant on a day-to-day basis. Exposures were determined as the mean exposure values per daily MCP or SDE. Third, time-weighted GPS-based buffers (TWB) with 100 m radii were created and superimposed on participants’ GPS points. This measure embodied all locations that any given participant visited during the tracking period. We selected 100 m buffers to capture an area that is immediately visible, or with which the respondent has direct contact ^24^. Time-weight per GPS point was calculated as the percentage of the time a participant spent at each GPS point of the total tracking duration. We determined the mean exposure estimates for each type of exposure measurement by obtaining time-weighted exposures by multiplying and summing exposure and time weight per GPS point.

#### 2.2.3 Refined GPS-based approach for context delineation

Figure 1 summarizes the four analytical steps to enable our geographic context delineation based on GPS data.

**Figure 1.**
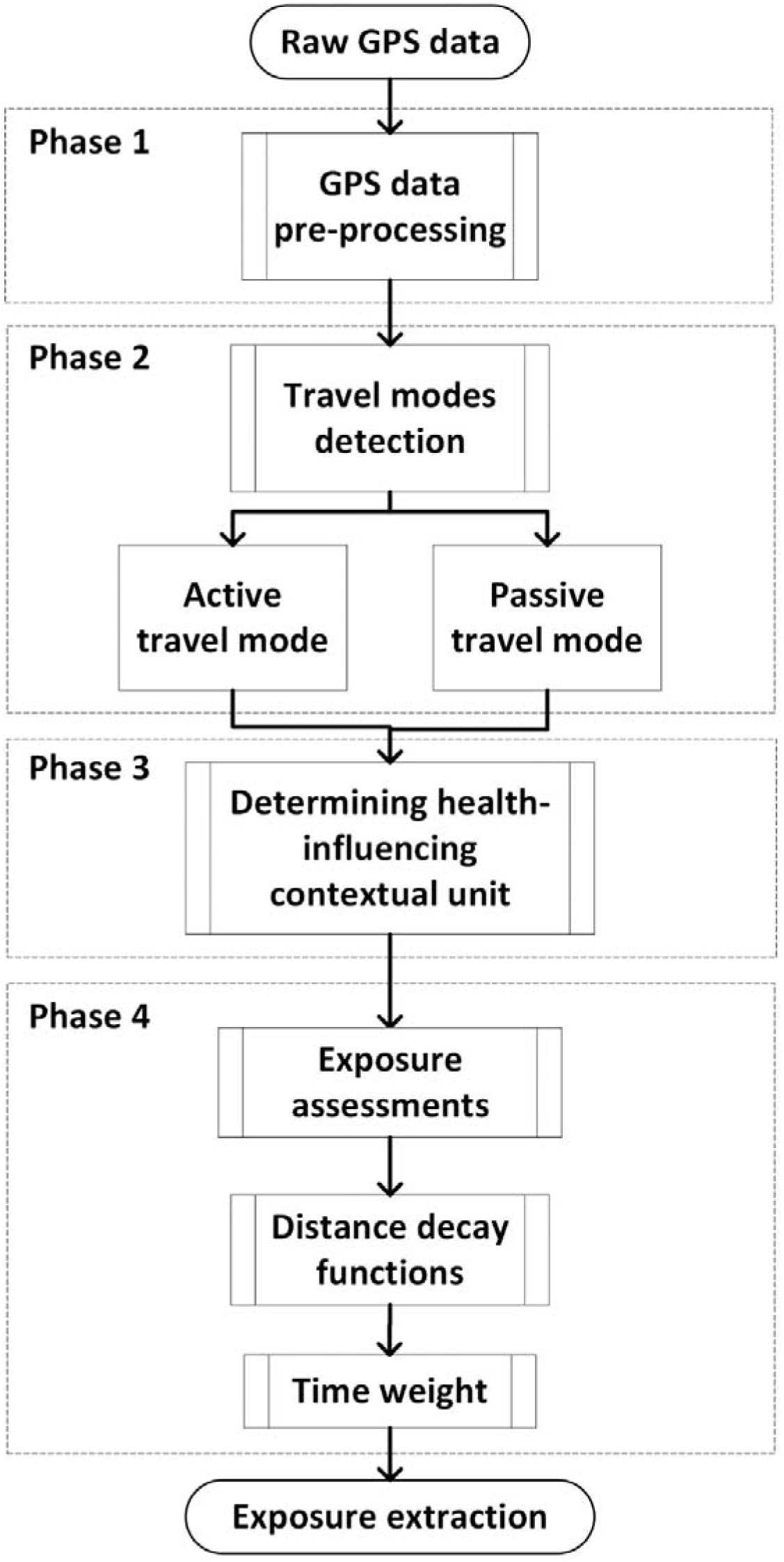
Workflow of the GPS-based refined context delineation approach.

##### Phase 1: GPS data pre-processing

The GPS data were cleaned in a three-step process to minimize the influence of anomalous records and increase data reliability. First, participants (*N*=188) with GPS records outside the Netherlands during the tracking period were excluded due to a lack of environmental data. Second, implausible data points were removed, including those with similar timestamps, records with a speed of >200 km/h, and a spatial displacement of >50 m away from the road network ^44^. These thresholds were selected based on Dutch traffic regulations and were aligned with previous GPS-based exposure assessments ^44, 45^. Third, we removed respondents (*N*=296) possessing insufficient GPS data points, defined as any number of data points less than two times the median absolute deviation of the number of GPS records ^46^. In total, 269 participants were retained after data pre-processing.

##### Phase 2: Travel modes detection

Before deriving travel modes, GPS data were segmented into individual trips as GPS points lack semantic information on mobility ^47^. Dwell time beyond a threshold, usually selected ad hoc, between consecutive GPS points is an established criterion for trip segmentation ^48^ but depends on the GPS sampling frequency, sensors, and urban setting ^49–51^. Based on previous studies ^45^, we set the dwell time threshold value at 3 minutes.

For travel mode detection (i.e., active travel mode and passive travel mode), previous studies used fuzzy systems ^52^, machine learning ^53^, or rule-based classifiers ^54^. However, model-based classifiers require pre-labeled training data for extracting feature characteristics of each travel mode, which were lacking in our case ^55^. As a result, we classified trips into active (e.g., walking, biking) and passive travel modes (e.g., car, bus) using traffic speed-based rules ^45^. To be classified as active, a trip was required to have an average trip speed of <25 km/h and a maximum speed of <45 km/h; otherwise, trips were labeled passive if they averaged less than 200 km/h. It is worth noting that some stationary GPS points around activity locations could also be labeled as active travel mode based on the implemented rules. Therefore, we referred to active travel mode and stationary locations as ’activity locations and active travel mode’.

##### Phase 3: Determining health-influencing contextual unit

In the case of passive travel mode, individuals are mainly exposed to the micro-environment in their vehicles and are less affected by perceptions of their extended surroundings ^35^. Consequently, buffer sizes for passive travel mode can be smaller than those for activity locations and active travel mode. Circular buffer sizes used for trips were set to 50 m and 100 m for passive travel mode and activity locations and active travel mode, respectively, as informed by previous studies ^56^.

##### Phase 4: Exposure assessments

Exposures were assessed in three steps. First, we extracted raster cells per environmental exposure within each buffer. Second, distance decay functions were used to approximate distance-dependent exposure influences. As assumed elsewhere ^19^, the exposure impacts on an individual declines with increasing distance. Different mechanisms of the distance decay exist for visible and invisible exposures. For visible exposures, the concentrations decrease with the increasing distance, while for invisible ones, the importance of locations varies with distance. To assess the sensitivity of the exposure assessment in terms of different distance decay functions, we implemented linear, exponential, and Gaussian functions (Fig. S2). In the linear case (LM), exposure weights constantly decreased with increasing distance. The exponential function (EM) captures a more rapid drop-off in weights within small distance increments, while in the case of the Gaussian function (GM) weights initially declined slowly and then more steeply as distance increased.

As a benchmark, we also implemented an unweighted model (UM) without any distance-based exposure weighting ^57^. The weighted exposure of each raster cell was calculated as the product of the distance-based weighting and the exposure values at the cell. Third, to consider the different dwell times people spent at different locations, we multiplied the time weight of each location by the corresponding weighted exposure and averaged the values across all GPS points.

### 2.3 Environmental data

#### 2.3.1 Green space

Green space was measured using the Normalized Difference Vegetation Index (NDVI) ^58^ with a 30 m spatial resolution derived from Landsat-8 satellite imagery. We obtained remote sensing images from Google Earth Engine ^59^ covering the Netherlands from May to September 2018. Images with cloud scores >25 and more than 40 percent cloud coverage were discarded. NDVI values range from -1 to 1, with negative values associated with less or no vegetation and positive values representing greater amounts of vegetation and, hence, green space. We excluded NDVI values <0 to avoid distortions when calculating the mean NDVI value per circular buffer.

#### 2.3.2 Noise

Estimated average noise data in day–evening–night noise levels (Lden) (in dB) for 24h periods emitted by roads (2017), rail, air traffic (2016), wind turbines (2020), and industrial areas were obtained from the Dutch National Institute for Public Health and Environment. Noise estimates with a spatial resolution of 10 m were derived from the Standard Model Instrumentation for Noise Assessments ^60^.

#### 2.3.3 Air pollution

Using land-use regression, nationwide average particulate concentrations with a diameter of <2.5 μm (PM_2.5_) (µg/m^3^) for the year 2016 were obtained with a 25 m spatial resolution. The land-use regression model was based on roads, land cover, and population data, among others, within concentric buffers of 50 to 10,000 m centered on the monitoring sites. Results of the model validation are provided elsewhere ^61^.

#### 2.3.4 Blue space

Blue space data were obtained from the Dutch land-use database (Landelijk Grondgebruiksbestand Nederland) for the year 2018 ^62^. With a spatial resolution of 5 m, the dataset classifies land use into forty-eight types. We extracted those raster cells classified as freshwater or saltwater to map blue space. Blue space exposure was measured as the proportion of blue space cells to the total number of cells in the buffer.

### 2.4 Statistical analysis

We used descriptive statistics (i.e., box plots) to compare the size of contextual units and the level of exposure across the activity space-based assessment methods. Because the exposures did not follow a normal distribution, we applied non-parametric statistical approaches to examine the statistical difference in size across different contextual units and environmental exposure levels. Kruskal-Wallis tests and additional post-hoc analysis using pair-wised Wilcoxon signed-rank tests were employed. We used pair-wise Spearman correlation analyses to assess the associations among exposures across each activity space-based exposure assessment. *P*-values were adjusted for multiple hypotheses testing ^63^. Analyses were performed in R, version 4.0.3 ^64^.

## 3 Results

### 3.1 Spatiotemporal characteristics of participants’ GPS points

The average time a person spent within and outside their residential neighborhoods (i.e., a 1 km buffer centered on the home location) per day was 13.2 hours (standard deviation (SD)±4.24) and 9.6 hours (SD±4.57), respectively (Table S1). Weekday versus weekend observations of the time spent within the residential neighborhood differed. People’s average distances for activities within and outside the residential neighborhood were 0.22 km (SD±0.11) and 14.74 km (SD±13.44), with negligible differences between weekdays and weekends. The SD implies that distances outside the residential neighborhood have greater variations across participants than those within the residential neighborhood. Greater deviations were observed on weekend days than weekdays.

**Table 1.** Median and interquartile range (IQR) of derived environmental exposures.

| Contextual unit | PM<sub>2.5</sub><br>($\mu\text{g}/\text{m}^3$) | | Noise<br>(dB) | | Green space<br>(NDVI) | | Blue space<br>(%) | |
| --- | --- | --- | --- | --- | --- | --- | --- | --- |
|  | Median | IQR | Median | IQR | Median | IQR | Median | IQR |
| HB | 12.83 | 1.92 | 55.91 | 6.84 | 0.43 | 0.12 | 0.03 | 0.05 |
| MCP | 12.58 | 1.52 | 54.81 | 6.07 | 0.45 | 0.09 | 0.06 | 0.06 |
| SDE | 12.23 | 1.59 | 52.99 | 6.46 | 0.45 | 0.07 | 0.07 | 0.07 |
| TWB | 12.89 | 1.63 | 56.75 | 6.24 | 0.38 | 0.11 | 0.02 | 0.04 |
| LM | 12.89 | 1.63 | 56.87 | 6.49 | 0.38 | 0.10 | 0.03 | 0.05 |
| EM | 12.89 | 1.63 | 56.83 | 6.28 | 0.38 | 0.10 | 0.03 | 0.05 |
| GM | 12.89 | 1.63 | 56.86 | 6.45 | 0.38 | 0.11 | 0.03 | 0.04 |
| UM | 12.89 | 1.63 | 56.87 | 6.29 | 0.38 | 0.10 | 0.03 | 0.05 |

Figure 2 shows the spatiotemporal density of participants’ GPS trajectory points over 24 hours. Not distinguishing between activity locations and active travel mode and passive travel mode, a higher density of GPS trajectory points was found for 10 to 100 km away from home between 7h and 16h and closer to home in the range of 0.1 km between 15h and 20h. Activity locations and active travel mode and passive travel mode exhibited distinct patterns. GPS trajectory points in activity locations and active travel mode clustered around people’s homes (around 0.1 km) between 14h and 22h. We observed dense passive travel mode GPS points between 10 km and 100 km during the morning and evening peak hours (i.e., between 6h and 8h, and around 16h).

**Figure 2.**
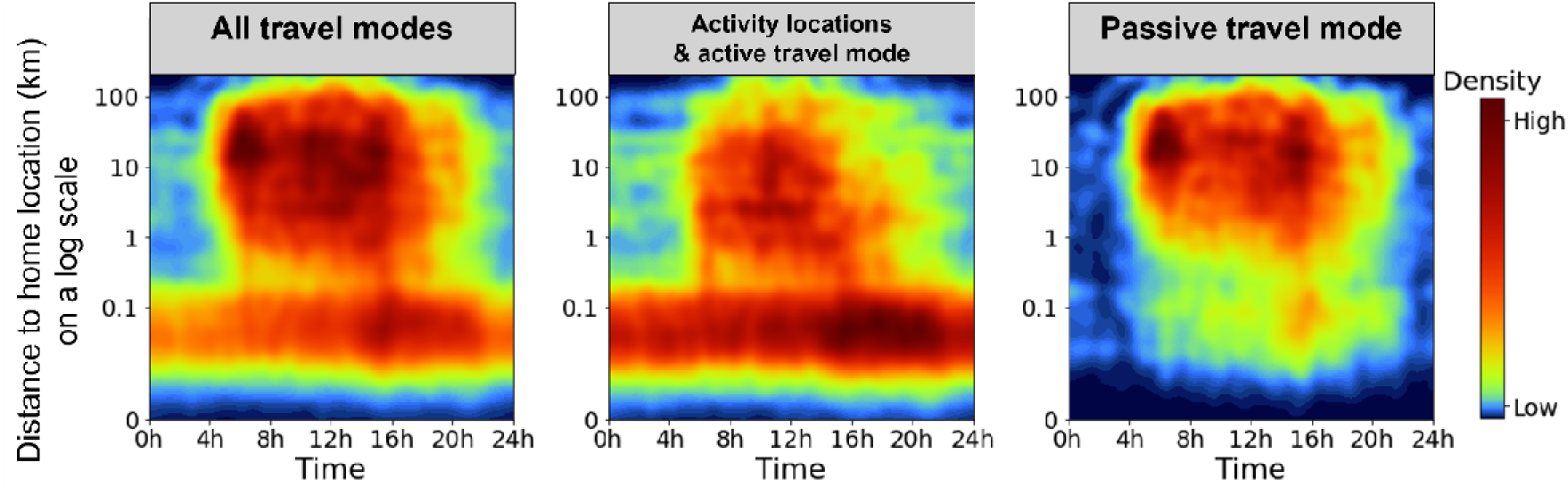
Spatiotemporal density of participants’ GPS trajectory points over 24h for each tracing day.

### 3.2 Comparison of the size of contextual units

Figure 3 compares the activity space-based context delineations. MCP and SDE showed a greater mean size than TWB and UM. SDE had the largest variance among the participants, and TWB had the smallest. Compared with TWB, UM had smaller maximum and minimum sizes but a greater variation among participants. Significant Kruskal-Wallis test results (Figure 3) and Wilcoxon test results (Table S2) showed that the sizes of the four contextual units were significantly different from each other (*p*<0.001). Additionally, all contextual units were larger than the size of HB.

**Figure 3.**
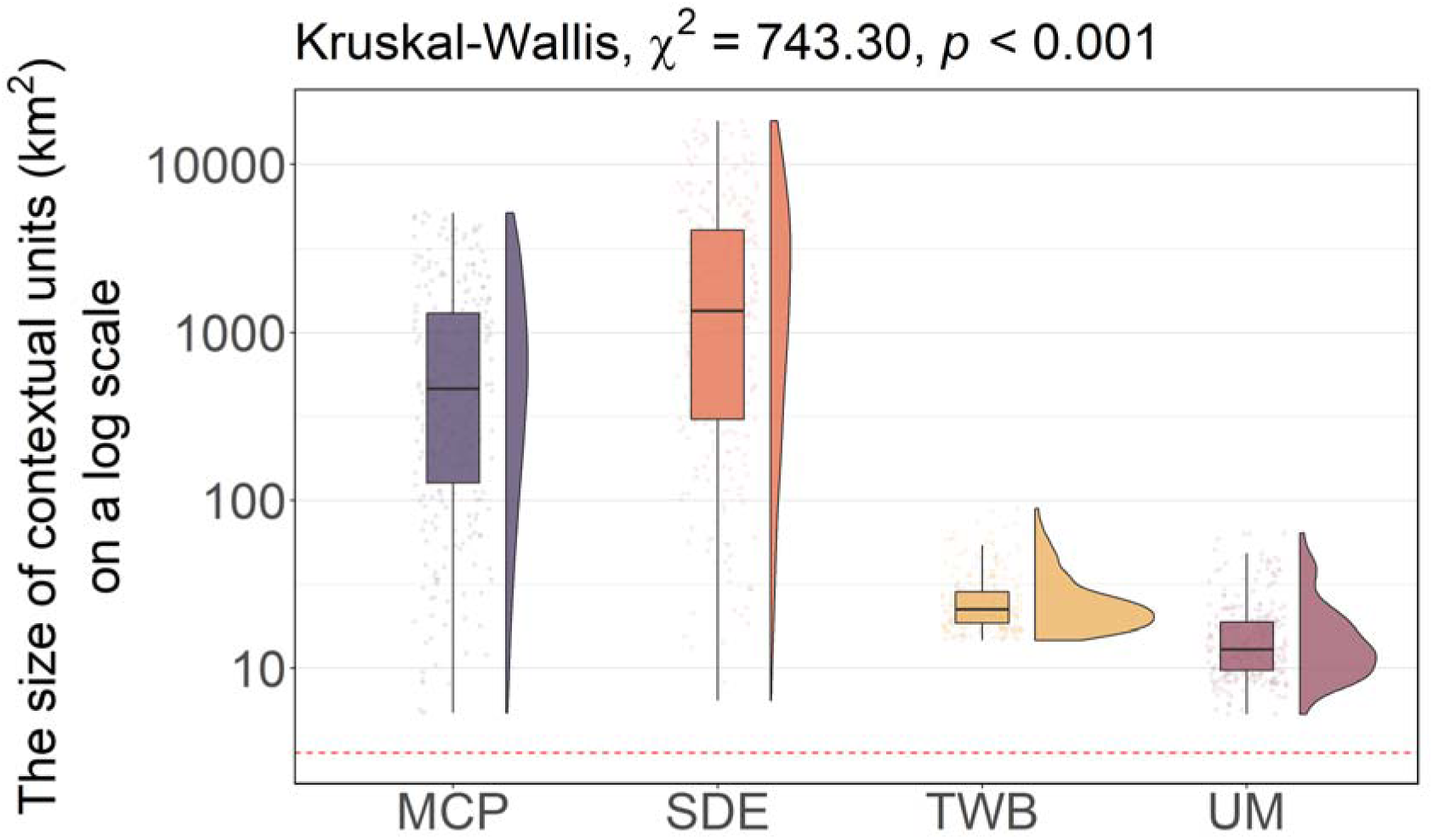
The size of different contextual units. The red dash line refers to the size of the 1 km home-based buffer (π ≈ 3.14 km^2^). Note that the 1 km home-based buffer was excluded from statistical examination because its size was identical for each participant. Since distance decay functions did not influence the sizes of contextual units, we report the unweighted model (UM) as the contextual unit that only includes travel modes.

### 3.3 Comparison of derived environmental exposures

Figure 4 and Table 1 summarize the exposure distributions of the eight tested activity space-based geographic context definitions. Home-based buffers (HB) showed large variations across all exposures except in the case of blue spaces. Conversely, MCP and SDE had the smallest interquartile range in PM_2.5_, noise, and green space but the largest in blue space. The minimum convex polygon (MCP) and two standard deviational ellipses (SDE) resulted in less pronounced PM_2.5_ and noise exposure levels; however, exposure to green and blue spaces was greater. The other contextual units exhibited similar exposure distributions and levels. EM and UM resulted in more compact noise distributions than LM and GM, while these models were similar in terms of the other exposures.

**Figure 4.**
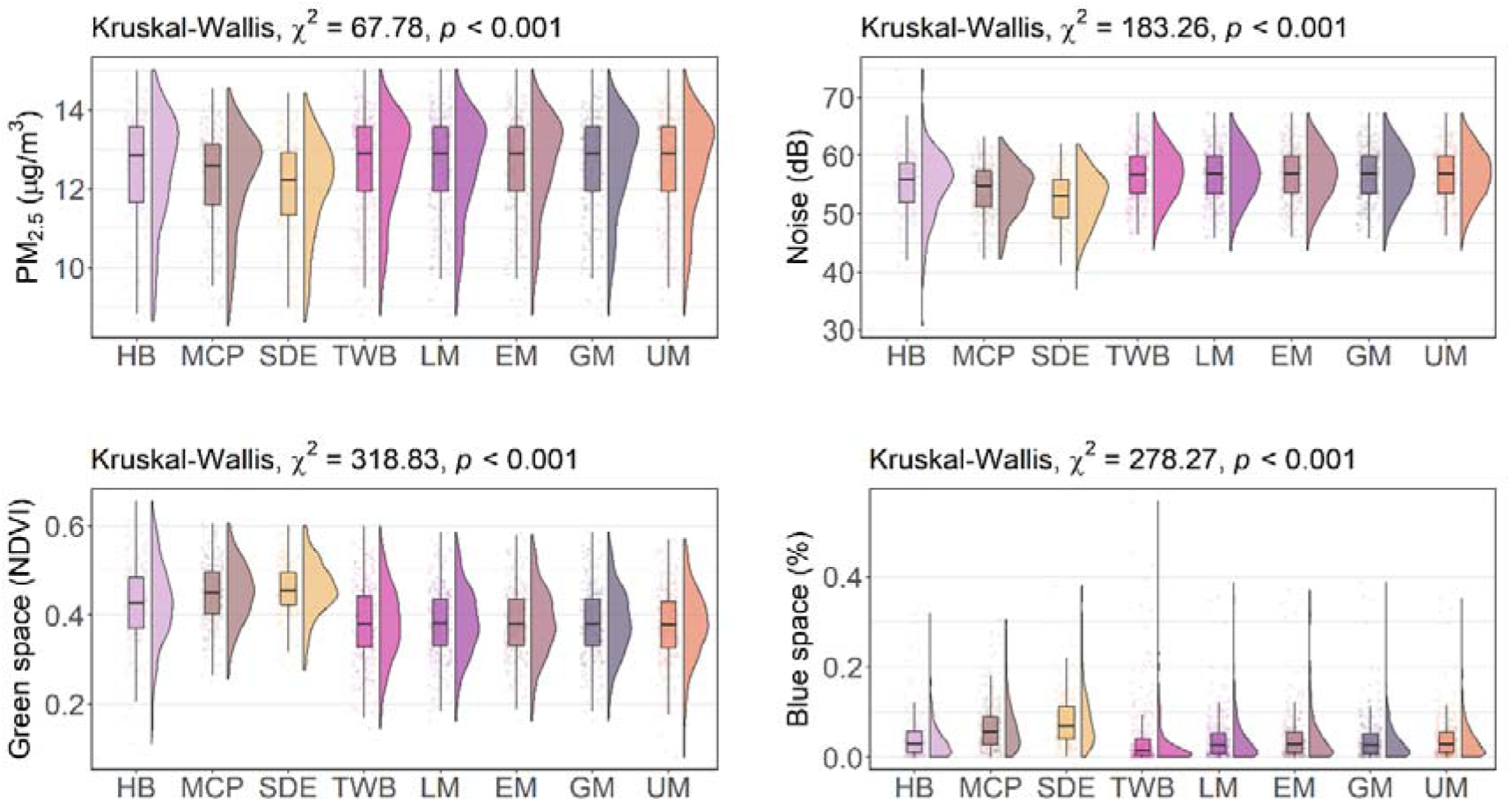
Boxplots of environmental exposures across different contextual units. Abbreviations: HB (home-based buffers); MCP (minimum convex polygon); SDE (two standard deviational ellipse); TWB (time-weighted GPS-based buffers); LM (linear model); EM (exponential model); GM (Gaussian model); UM (unweighted model).

The Kruskal-Wallis tests were significant for all exposure assessments (*p*<0.001). Wilcoxon tests showed that the linear model (LM) versus Gaussian model (GM) test pair was the only significant test pair across four distance decay models in PM_2.5_ assessments (see Table S3). Concerning noise, the linear model (LM) was not significantly different from the Gaussian model (GM), and no statistical differences were obtained between the time-weighted GPS-based buffer (TWB) and four distance decay models. For green space, the results indicated no statistical difference in the following test pairs: time-weighted GPS-based buffer (TWB) versus unweighted model (UM), linear model (LM) versus exponential model (EM), exponential model (EM) versus Gaussian model (GM), and linear model (LM) versus Gaussian model (GM). In the case of blue space, the linear model (LM), exponential model (EM), Gaussian model (GM), and unweighted model (UM) were all insignificantly different from the home-based buffer (HB). We also found no statistical differences across the linear model (LM), exponential model (EM), Gaussian model (GM), and unweighted model (UM).

The Spearman correlations are depicted in Figure 5. The results show that exposure estimates across different contextual units were significantly correlated and all estimated PM_2.5_ exposure levels were highly correlated (0.87 - 1). As for other exposures, the minimum convex polygon, two standard deviational ellipse, and home-based buffers showed only moderate correlations with the time-weighted GPS-based buffer, unweighted model, linear model, exponential model, and Gaussian model. Correlations between the unweighted, linear, exponential, and Gaussian models in assessing all exposures were strong (0.94 - 1).

**Figure 5.**
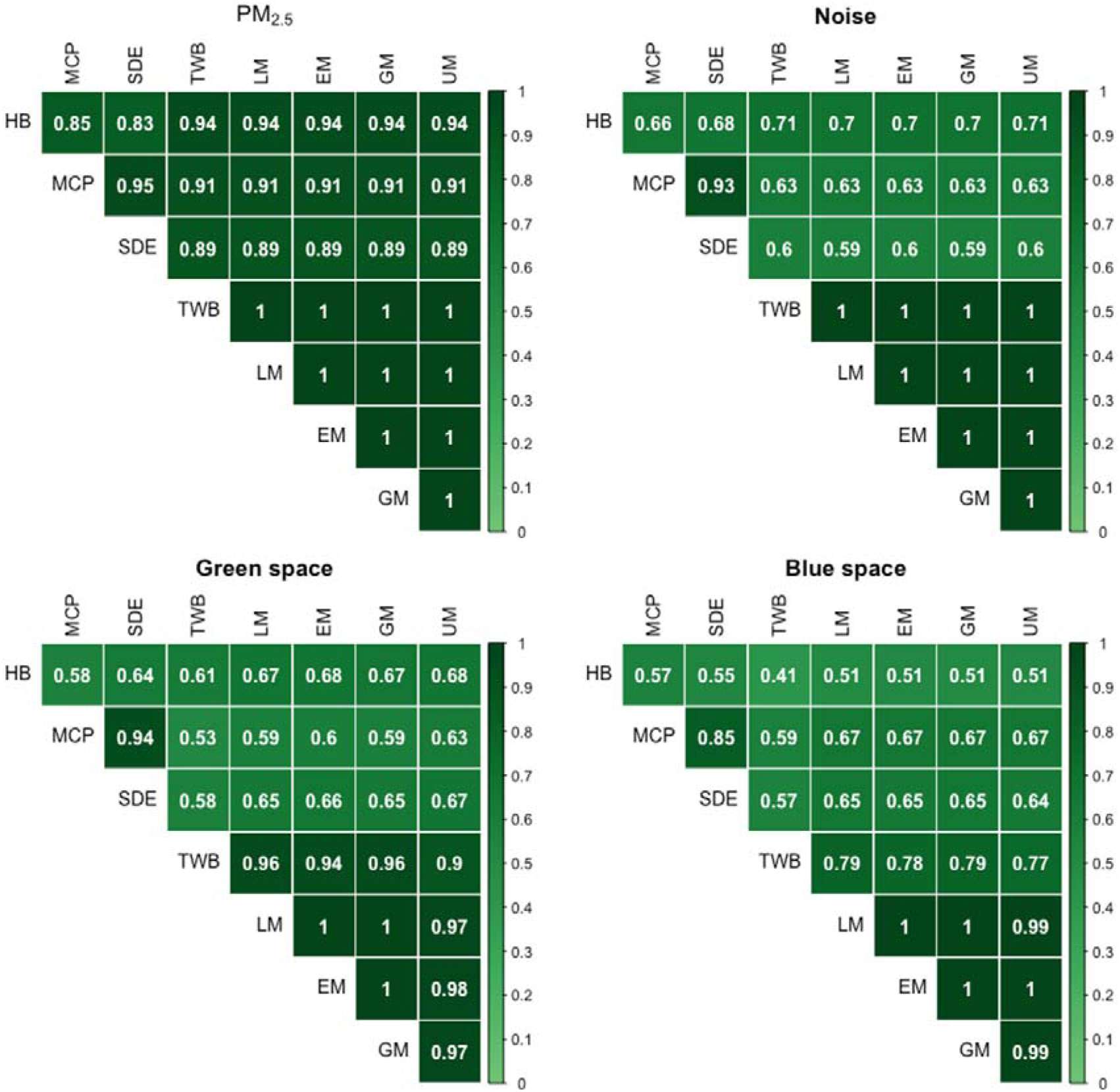
Spearman correlation matrices of environmental exposures across different contextual units. Abbreviations: HB (home-based buffers); MCP (minimum convex polygon); SDE (two standard deviational ellipse); TWB (time-weighted GPS-based buffers); LM (linear model); EM (exponential model); GM (Gaussian model); UM (unweighted model).

## 4 Discussion

This study extended activity space-based exposure assessments by accounting for travel modes and exposure-related distance decay effects to allow evaluation of the size of activity spaces and exposure levels against contextual units in typical use.

### 4.1 Main findings

In line with our expectations, our results showed that people’s daily mobility extends beyond their residential neighborhoods. Including travel modes in the exposure assessments significantly reduced contextual unit sizes confirming our first hypothesis. However, our first hypothesis about assessed exposure levels had to be rejected. Significant statistical differences were only found for PM_2.5_ and blue space for cases with or without considering travel modes in exposure assessments. We found that including travel modes in the assessment decreased measured exposure levels for most participants. Our second hypothesis was partially supported. We observed that incorporating distance decay functions was sensitive to the type of exposures. For PM_2.5_ and blue space, no statistical differences were found between the unweighted model (UM) and linear (LM), exponential (EM), and Gaussian (GM) models. By contrast, differences in distance decay models appeared to be significant for noise and green space. Our results reported insignificant differences in derived exposure levels across multiple distance decay functions.

The high correlation of 0.94 between home-based buffers (HB) and time-weighted GPS-based buffers (TWB) suggested that the amount of PM_2.5_ measured around residential locations is an excellent proxy for the exposure experienced during daily life in the Netherlands. For other exposures (i.e., noise, green space, and blue space), HB can only moderately approximate daily exposure which implies that GPS point-based assessments are more suitable in such cases. Moreover, TWB provides a good estimate of noise and green space exposure, as evidenced by the highly correlated relationships between TWB and LM, EM, and GM, and UM (0.9 < correlation coefficients <1). TWB and UM were potentially excellent proxies for the overall blue space exposure along individuals’ daily mobility paths.

### 4.2 Available evidence

Our findings support prior studies suggesting that human spatiotemporal activity patterns should be incorporated in exposure assessment ^32, 65, 66^. Furthermore, the results comparing environmental exposure levels captured with different activity space-based approaches were congruent with past research ^24, 67, 68^. The results underscore that contextual uncertainty is critical in dynamic exposure assessment as different delineations of activity space exhibit significant discrepancies in observed levels of exposure ^19, 20, 69^.

We found that the minimum convex polygon (MCP) and two standard deviational ellipses (SDE) resulted in lower exposure concentrations for PM_2.5_, lower noise exposure, and higher exposure to green space and blue space than other contextual units. In this context, sparsely distributed GPS points versus clustered points ^20^ could lead to much larger contextual unit sizes. Such contextual units may include spaces (e.g., parks) an individual has never visited and thus lead to incorrect estimates of actual exposure. Consequently, as reported in past studies, MCP and SDE are not suitable contextual units for exposure estimates ^19^.

Home-based buffers (HB) showed large variations across participants for PM_2.5_ and green space but fewer variations for noise and blue space. Significant PM_2.5_ and green space variations could be attributed to participants’ socioeconomic disparities. Lower socioeconomic status has been observed to relate to living conditions associated with higher air pollution levels and a lower presence of green space ^70, 71^. Exposures to noise and blue space are less likely to be affected by participants’ socioeconomic status. However, the evidence about whether noise exposure differs across socioeconomic groups is mixed ^72^. In the case of noise, this may be because loud noise may lead to anxiety and sleep disturbance ^73^, causing people to avoid choosing a noisy living environment. In the case of blue space, the possible reason is that the percentage of blue space is somewhat limited within the 1 km radius of the living neighborhoods we examined.

We also observed that including travel modes significantly influenced the exposure assessments. This observation may be due to selective daily mobility bias, potentially affecting individuals’ choices and attitudes toward travel ^74^. People tend to choose travel-friendly routes given choices in travel pattern creation ^75^ and ambient exposure levels^76^. In terms of exposure to PM_2.5_, traffic emissions have been recognized as one of the primary sources. Past research has observed high levels of exposure along transportation routes ^34, 77^; thus, commuters can attenuate their exposure by minimizing the use of such routes, increasing their distance from emission sources ^78^. In the case of passive travel mode, individuals’ mobility paths are co-located with the emitting paths, increasing their exposure levels. By contrast, active travelers can often choose less-polluted routes improving their travel environments ^79^. Finally, in the case of blue space, a possible explanation is that blue space could promote walking and cycling activities ^80, 81^ while also constituting a barrier to vehicular traffic.

The significant impact of the distance decay effect on exposure assessment only held for noise and green space. Our inconclusive results concerning blue space may be attributed to its limited presence and measurement in our data. First, the presence of blue space only accounts for a small percentage of individuals’ activity space, especially considering people’s mobility paths. Second, we used the buffers’ average proportion of blue space as a proxy for blue space exposure. However, such a means of measurement may be insensitive to the distance decay effect. As for PM_2.5_, the nature of exposure might be a possible explanation for the insignificant role of the distance decay effect. Visible exposures might affect human health within limited geographical areas, and such effects decrease with distance ^82, 83^. By contrast, invisible presence (i.e., PM_2.5_) might be less sensitive to distance. Compared with previous studies ^19^, we cannot conclude which distance decay function performs better or worse. Nonetheless, a novel contribution was made to the literature as the first study to consider the distance decay effect in measuring environmental exposures.

Surprisingly, residential PM_2.5_ exposure (HB) correlated highly with other activity space-based exposure estimates. Even though a prior study in the Netherlands noted that residential exposure was sufficient to capture school children’s annual average PM_2.5_ exposure ^68^, such a finding was at variance with other existing findings and still not expected. The finding might be accounted for by differences in the geographic context of the Netherlands compared to other contexts ^84^. Despite participants spending a great deal of time outside their homes, residential exposure would appear to be an excellent proxy for the overall exposure to PM_2.5_ in daily life in the Netherlands, likely caused by the spatial disparity of PM_2.5_ concentrations in the Netherlands not being significant. As a result, studies lacking GPS data availability could still adopt residential exposure as a reliable proxy value for assessing PM_2.5_ exposure in the Netherlands.

In contrast with PM_2.5_ exposures, residential exposure values were not suitable as proxy values for other environmental exposures encountered over individuals’ daily mobility. Such exposures are best assessed using GPS-based buffers. The strong correlations between time-weighted GPS-based buffers (TWB) and other distance decay models for noise and green space assessments suggest that TWB is a less-costly and more straightforward GPS data-based approach. In the case of blue space, either including travel modes in exposure assessment or using TWB are both potentially suitable approaches. We recommend that the selection of exposure assessment approaches be approached with care and informed by applying sensitivity tests and examining the actual data.

### 4.3 Strengths and limitations

Among the first, this study systematically compared people’s exposure to air pollution, noise, green space, and blue space across different activity space-based contextual units advanced on previous studies, which primarily assessed home-centered exposures ^13, 14^. Benefiting from GPS data, multiple dynamic activity space-based contextual units were delineated. As our participants’ mobility trajectories were distributed across the Netherlands, this enabled our recommendations about the performance of contextual units in different exposure assessments to be adaptive to different environmental settings throughout the country. Finally, we extended current research practice by refining the GPS-based assessment of environmental exposures allowing for accounting of travel modes and exposure-based distance decay functions. However, selecting a specific distance decay function will require further investigation and considering actual needs.

This study was subject to several limitations. First, the GPS data did not contain semantic information about people’s travel modes. We cannot exclude that our rules-based classification has affected the classification accuracy to some extent. Second, the GPS data quality could be affected by the signal reception (e.g., high buildings, tree canopy) and data pre-processing. Concerning the latter, we cannot rule out that a few meaningful GPS points were erroneously removed despite satisfying the elimination criteria. However, since only 3.7% of the GPS points were removed, the impact is likely neglectable. Similarly, the amount of problematic GPS points (e.g., points falling clearly off the potential path) was also negligible. Third, our data only focused on short-term environmental exposures. The statistical differences across different activity space-based approaches in short-term exposures could cumulatively contribute to substantial variations in long-term exposure levels ^69^. Furthermore, an investigation is needed to assess the differences between short- and long-term exposures based on different contextual units. Fourth, our study ignored short-term temporal variation in environmental exposures as typically done in GPS studies ^27, 28^. Relatedly, there was also a temporal mismatch between the GPS data collection and the environmental exposure data.

Such a mismatch could potentially affect the exposure levels. Fifth, we applied the same distance decay functions across exposures, which could potentially influence the results. Sixth, the smartphone app for collecting GPS data was unavailable for the iOS system; thus only Android system users could be approached and recorded. Even though the Android system accounted for 76.3% of the Dutch market at the time of data collection, specific groups of people could potentially be excluded.

## 5 Conclusions

Our results showed that for some exposures, the tested activity space definitions, although significantly correlated, exhibited differing exposure estimate results based on the inclusion or exclusion of travel modes or distance decay effects. Furthermore, residential exposure appears to be an excellent proxy for the overall amount of PM_2.5_ exposure in daily life, with GPS point-based assessment suitable for other exposures.

## Supporting information

see supplementary Fig. S1

## Data Availability

The GPS data used in this analysis cannot be shared with third parties as per the policies of Statistics Netherlands.

## Author contributions

*Lai Wei*: Conceptualization, Methodology, Formal analysis, Visualization, Writing – original manuscript. *Mei-Po Kwan*: Writing – review & editing, Funding acquisition. *Roel Vermeulen*: Funding acquisition, Project administration. *Marco Helbich*: Conceptualization, Data Curation, Methodology, Writing – review & editing, Supervision, Funding acquisition.

## Declaration of interest

None.

## Conflict of interest

The authors have no conflict of interest to declare.

## Funding

This work received funding from the European Research Council (ERC) under the European Union’s Horizon 2020 research and innovation program (grant agreement No 714993). The study was supported by EXPOSOME-NL, which is funded through the Gravitation program of the Dutch Ministry of Education, Culture, and Science and the Netherlands Organization for Scientific Research (NWO grant number 024.004.017).

## Footnotes

1 The contextual unit represents the geographical area (in km^2^) used as an analytical unit when examining the effects of area-based exposures on individual-level outcomes.

## Notes

### Competing Interest Statement

The authors have declared no competing interest.

### Author Declarations

The Ethics Review Board of Utrecht University gave ethical approval for this work.

