## Supplementary material for "Measuring environmental exposures in people’s activity space: The need to account for travel modes and exposure decay": see supplementary Fig. S1

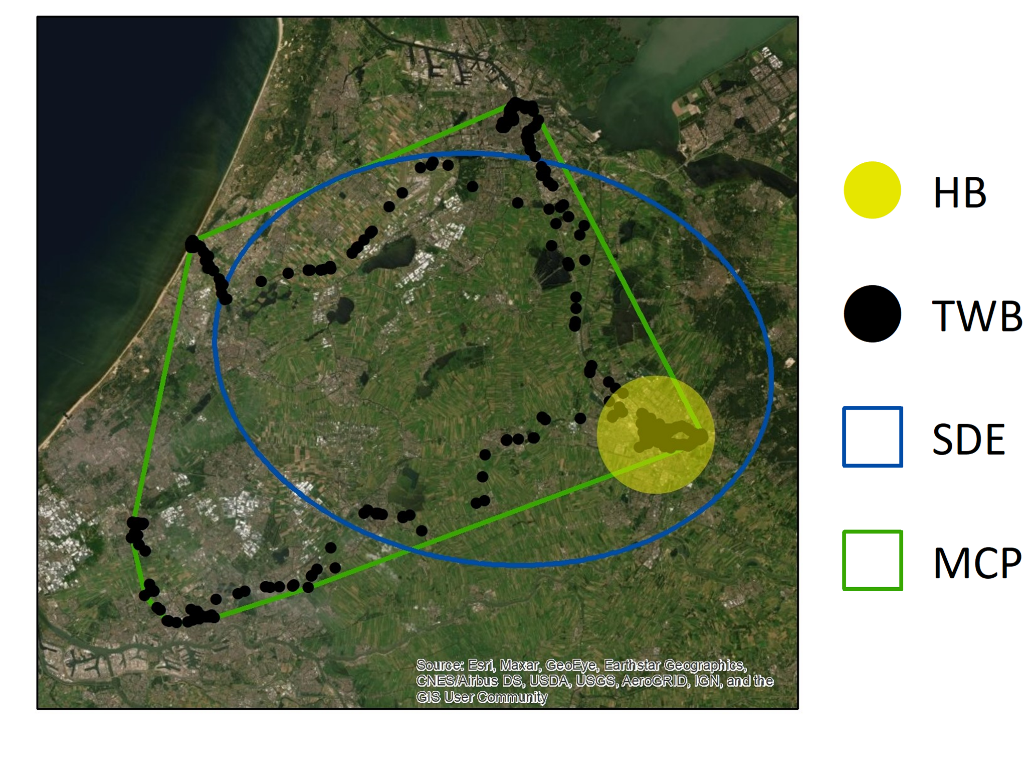

**Figure S1**. Visualization of four alternative contextual units (For privacy protection, this figure is drawn based on test data, and detailed geographic information is erased). Abbreviations: HB (home-based buffer); TWB (time-weighted GPS-based buffer); SDE (two standard deviational ellipse); MCP (minimum convex polygon). Source: Esri, Maxar, GeoEye, Earthstar Geographics, CNES/Airbus DS, USDA, USGS, AeroGRID, IGN, and the GIS User Community.

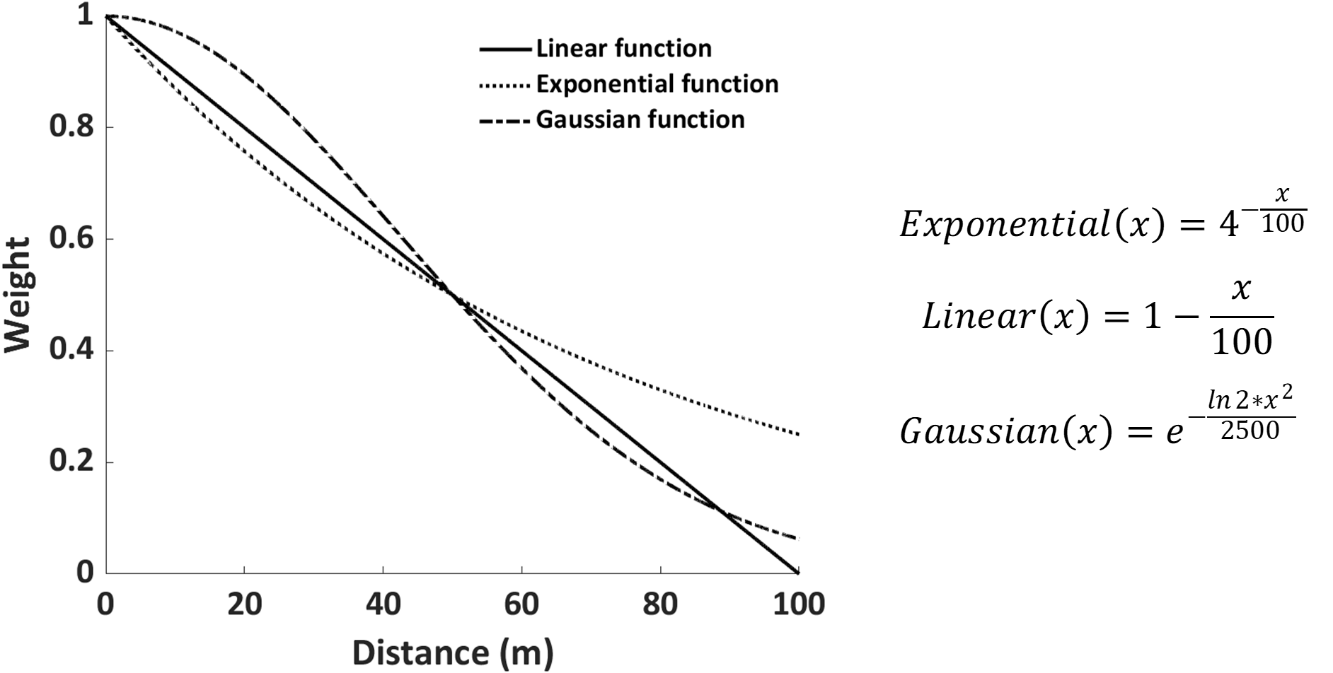

**Figure S2**. Three classic distance decay functions.

**Table S1.** Spatiotemporal characteristics of participants' GPS trajectory points within and outside their residential neighborhoods (Residential neighborhood is defined as the neighborhood within 1 km of home location).

|  | **Distance to home**  **Mean (SD)** | | **Duration**  **Mean (SD)** | |
| --- | --- | --- | --- | --- |
|  | Within residential neighborhood | Outside residential neighborhood | Within residential neighborhood | Outside residential neighborhood |
| **Weekdays** | 0.23 km (0.12) | 14.80 km (13.89) | 12.98 hours (4.38) | 9.64 hours (4.66) |
| **Weekends** | 0.20 km (0.14) | 15.06 km (20.92) | 15.22 hours (7.85) | 10.05 hours (8.17) |
| **All-day** | 0.22 km (0.11) | 14.74 km (13.44) | 13.20 hours (4.24) | 9.60 hours (4.57) |

**Table. S2**. Wilcoxon signed-rank test of the size of different contextual units.

|  | *MCP* | *SDE* | *TWB* |
| --- | --- | --- | --- |
| *SDE* | -14.157^a**^ |  |  |
| *TWB* | -13.945^b**^ | -14.147^b**^ |  |
| *UM* | -14.032^b**^ | -14.164^b**^ | -14.217^b**^ |

** = p < 0.01.

^a^. Based on negative ranks.

^b^. Based on positive ranks.

**Abbreviations:** MCP (minimum convex polygon); SDE (two standard deviational ellipse); TWB (time-weighted GPS-based buffer); UM (unweighted model)

| **Table. S3.** Wilcoxon signed rank tests of environmental exposures across different | | | | | | | |
| --- | --- | --- | --- | --- | --- | --- | --- |
| contextual units. | |  |  |  |  |  |  |
| **PM_2.5_** | *HB* | *MCP* | *SDE* | *TWB* | *LM* | *EM* | *GM* |
| *MCP* | -6.443^b**^ |  |  |  |  |  |  |
| *SDE* | -10.741^b**^ | -11.180^a**^ |  |  |  |  |  |
| *TWB* | -4.843^b**^ | -11.007^b**^ | -13.505^b**^ |  |  |  |  |
| *LM* | -4.721^b**^ | -10.972^b**^ | -13.487^b**^ | -2.597^a**^ |  |  |  |
| *EM* | -4.756^b**^ | -10.986^b**^ | -13.495^b**^ | -3.027^a**^ | -1.186^b^ |  |  |
| *GM* | -4.709^b**^ | -10.973^b**^ | -13.487^b**^ | -2.599^a**^ | -2.250^a*^ | -1.324^a^ |  |
| *UM* | -4.799^b**^ | -10.991^b**^ | -13.501^b**^ | -3.861^a**^ | -1.310^a^ | -1.480^a^ | -1.371^a^ |
| **Noise** | *HB* | *MCP* | *SDE* | *TWB* | *LM* | *EM* | *GM* |
| *MCP* | -3.245^b**^ |  |  |  |  |  |  |
| *SDE* | -8.781^b**^ | -12.621^a**^ |  |  |  |  |  |
| *TWB* | -6.261^b**^ | -9.404^b**^ | -12.324^b**^ |  |  |  |  |
| *LM* | -6.093^b**^ | -9.254^b**^ | -12.195^b**^ | -1.530^a^ |  |  |  |
| *EM* | -6.188^b**^ | -9.338^b**^ | -12.274^b**^ | -0.384^a^ | -3.469^b**^ |  |  |
| *GM* | -6.085^b**^ | -9.238^b**^ | -12.182^b**^ | -1.554^a^ | -0.926^a^ | -3.391^a**^ |  |
| *UM* | -6.292^b**^ | -9.444^b**^ | -12.327^b**^ | -1.010^b^ | -3.462^a**^ | -3.379^a**^ | -3.404^a**^ |
| **Green space** | *HB* | *MCP* | *SDE* | *TWB* | *LM* | *EM* | *GM* |
| *MCP* | -4.470^a**^ |  |  |  |  |  |  |
| *SDE* | -6,722^a**^ | -6.124^b**^ |  |  |  |  |  |
| *TWB* | -8.572^a**^ | -11.635^a**^ | -12.624^a**^ |  |  |  |  |
| *LM* | -8.892^a**^ | -12.233^a**^ | -13.252^a**^ | -2.004^b*^ |  |  |  |
| *EM* | -8.986^a**^ | -12.376^a**^ | -13.364^a**^ | -1.965^b*^ | -1.340^b^ |  |  |
| *GM* | -8.882^a**^ | -12.209^a**^ | -13.237^a**^ | -2.033^b*^ | -0.958^a^ | -1.332^a^ |  |
| *UM* | -9.698^a**^ | -12.826^a**^ | -13.597^a**^ | -0.230^a^ | -2.598^b**^ | -2.900^b**^ | -2.576^b*^ |
| **Blue space** | *HB* | *MCP* | *SDE* | *TWB* | *LM* | *EM* | *GM* |
| *MCP* | -8.647^a**^ |  |  |  |  |  |  |
| *SDE* | -10.900^a**^ | -7.221^b**^ |  |  |  |  |  |
| *TWB* | -3.207^a**^ | -8.915^a**^ | -10.416^a**^ |  |  |  |  |
| *LM* | -0.097^a^ | -9.025^a**^ | -11.809^a**^ | -3.549^b**^ |  |  |  |
| *EM* | -0.183^b^ | -9.042^a**^ | -11.812^a**^ | -3.579^b**^ | -1.441^b^ |  |  |
| *GM* | -0.118^a^ | -9.011^a**^ | -11.806^a**^ | -3.597^b**^ | -1.237^b^ | -1.325^a^ |  |
| *UM* | -0.426^b^ | -9.009^a**^ | -11.731^a**^ | -3.512^b**^ | -1.564^a^ | -1.527^a^ | -1.430^a^ |
| * = *p* < 0.05; ** = *p* < 0.01. | | |  |  |  |  |  |
| ^a^. Based on positive ranks. | |  |  |  |  |  |  |
| ^b^. Based on negative ranks. | |  |  |  |  |  |  |
| **Abbreviations:** HB (home-based buffers); MCP (minimum convex polygon); SDE (two standard deviational ellipse); TWB (time-weighted GPS-based buffers); LM (linear model); EM (exponential model); GM (Gaussian model); UM (unweighted model). | | | | | | |  |
